# Quality and Utilisation of DHIS2 data for health decision making and advocacy in Kenya: A Qualitative Study

**DOI:** 10.1101/2025.03.25.25324643

**Authors:** Phoene Mesa Oware, Gregory Omondi, Celestine Adipo, Mohamed Adow, Conrad Wanyama, Dan Odallo, Nelly Bosire, Lawrence Okong’o, Michuki Maina, Jalemba Aluvaala, Peter Ngwatu, David Githanga, Doris Kinuthia, Irene Amadi, Grace Rukwaro, Linet Kerubo, Cynthia Amisi, Allan Govoga, Janette Karimi, Ali Kahtra, Andrew Mulwa, Patrick Amoth, Ambrose Agweyu, Fred Were

**Affiliations:** Kenya Paediatric Research Consortium, Nairobi, Kenya; School of Public Health, University of the Western Cape, Cape Town, South Africa; KEMRI-Wellcome Trust Research Programme, Nairobi, Kenya; Kenya Paediatric Association, Nairobi, Kenya; University of Nairobi, Nairobi, Kenya; Ministry of Health, Nairobi, Kenya; Council of Governors, Nairobi, Kenya; London School of Hygiene and Tropical Medicine, London, UK

**Keywords:** District Health Information Software 2 (DHIS2), Health Information Systems, Kenya, Primary Health Care (PHC), Reproductive, Maternal, New-born, Child, Adolescent Health and Nutrition RMNCAH+N)

## Abstract

Reliable health information systems (HIS) are critical for effective decision-making in the delivery of Primary Health Care and Reproductive, Maternal, Newborn, Child, Adolescent Health and Nutrition (PHC/RMNCAH+N) services. In Kenya, the District Health Information Software 2 (DHIS2) platform serves as the primary HIS for tracking health indicators. This study explored perceptions of DHIS2 data quality and use for decision-making among PHC/RMNCAH+N stakeholders across 15 counties in Kenya. A qualitative study design was used, incorporating interviews with 88 key informants who were PHC/RMNCAH+N stakeholders in 15 counties in Kenya, to explore experiences, barriers, and facilitators of DHIS2 data use. Thematic network analysis was employed to identify recurrent themes and generate insights into the utility of DHIS2-generated information. Sociotechnical challenges included limited technical capacity among health staff, inadequate analytical skills, and reliance on a small pool of Health Records Information Officers (HRIOs), which hindered effective use of DHIS2 data. However, positive practices emerged, such as the use of DHIS2 dashboards and user-friendly outputs, which were valued for supporting evidence-based decision-making and advocacy, particularly at higher levels of health management. In some counties, visual displays of data, including scorecards and performance trends, facilitated budget advocacy and community engagement. Contextual barriers, such as disparities in access to data collection tools post-devolution, human resource shortages, and limited integration of private sector data, contributed to incomplete reporting. These challenges underpinned perceived inaccuracy of DHIS2 data, hindering the complete reliance on DHIS2 data for planning and decision making. The study highlights the need for targeted investments to improve DHIS2 data quality and use through stronger stakeholder coordination, enhanced data synthesis skills, and fostering a culture of data ownership both at the county and health facility levels. Addressing these gaps will contribute to improvement in DHIS2 data quality, enhanced ownership and reliance on DHIS2 data by PHC/RMNCAH+N stakeholders for decision making in Kenya.

## Introduction

Health Information Systems (HIS) are essential components of a health system, integrating data collection, processing, reporting, and use of health information to enhance efficiency and effectiveness of services (1). A well-functioning HIS enables decision makers at all levels to identify challenges, engage in evidence-based decision-making, and optimize the allocation of limited resources (2). Strengthening HIS is thus essential for tracking progress towards health goals, including Sustainable Development Goal (SDG) 3 – good health and wellbeing (3,4).

Despite the importance of robust HIS, concerns remain over their capacity in many low- and middle-income countries (LMICs), where health systems often face challenges due to fragmented information landscapes, inadequate infrastructure, and limited resources (2,3). In response to these challenges, significant investments have been made to improve HIS. These efforts have often focused on enhancing the capacity of the District Health Information Software 2 (DHIS2), an open-source platform widely adopted for managing health data (5). DHIS2 is widely adopted and is in use by at least 80 LMICs, at national and sub-national levels (5). It can be customized to meet specific country/organizational needs; it is supported by basic hardware; and it offers easy analysis and real-time data visualization (6).

Kenya adopted and rolled out the DHIS2 system software nationwide in 2011 (7). This coincided with a transition to a devolved system of governance, including health governance (7). The previous structure, with 8 administrative units (provinces), was replaced with 47 administrative units (counties) (8). In the devolved system, county governments are responsible for overseeing health services, including public health facilities that are organized into 4 levels: community services (level 1), dispensaries and clinics (level 2), health centres, maternity and nursing homes (level 3), and county referral services (level 4) (9). The national government retained critical functions such as policy formulation, quality and standards control, and the management of national referral institutions (level 5) (9). The devolution of health governance created a demand for data by county health managers who need to utilize DHIS2 generated information for decision-making including planning, budgeting, and monitoring programs, as well as among local elected leaders who rely on data to support their political manifestos and re-election bids (7).

While the technical and operational elements of DHIS2 are relatively well understood (6,10), it is not clear to what extent information generated via the platform is being used for decision making in LMICS (11).

### A socio-technical approach to HIS data use

A social technical perspective (Figure 1) to use of technology e.g., DHIS2 software in organizations, e.g., health management institutions posits that outcomes, such as evidence-based decision making, result from interdependent relationships between social/behavioural and technical sub-systems within an organization (12). The social/behavioural sub-system (comprising elements such as members of an organization and relationships among them, organizational values, culture, goals) are recursively and reciprocally related to the technical sub-system (e.g., technologies such as DHIS2 software, techniques and skills required to operate them) (12,13). As such, optimizing one sub-system alone, social/behavioural, or technical, without consideration for the other, results in the sub-optimization of the other or the socio-technical whole (14). Optimal outcomes are realized when a ‘goodness of fit’ is achieved between social and technical elements within an organization (15). The social and technical subsystems are however engulfed and are influenced by the environmental subsystem which is the political, economic and social context within which the social and technical sub systems operate (13).

**Figure 1.**
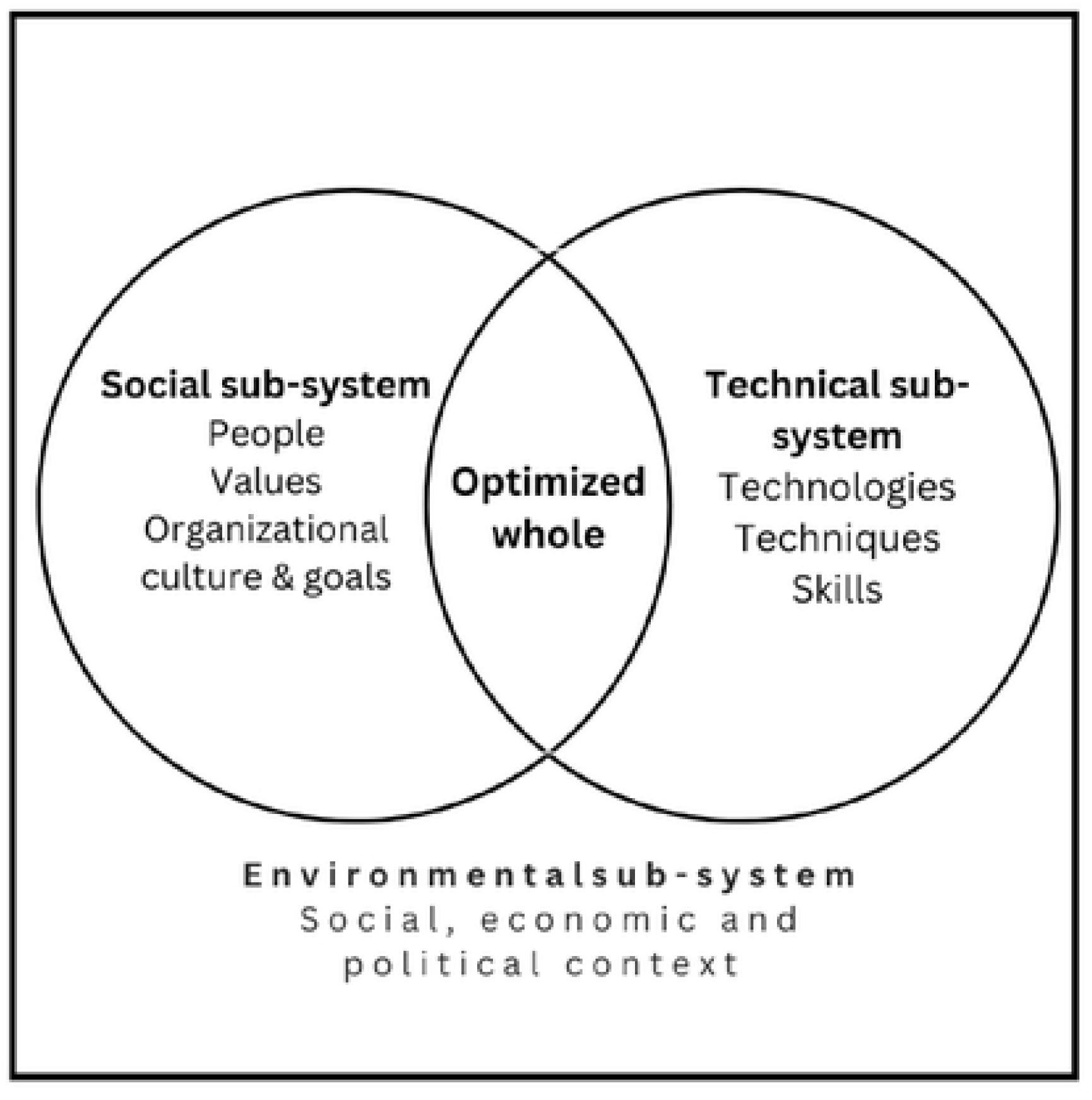
Dimensions of a socio-technical sub-system. Adapted from Kaminski J. Editorial. Theory applied to informatics Socio-technical theory. Canadian Journal of Nursing Informatics. 2022;17(3–4).

We draw on this socio-technical perspective to study the perceptions of DHIS2 data quality and its use by health decision makers to improve Primary Health Care (PHC) services with a focus on reproductive maternal child and adolescent health and nutrition (PHC/RMNCAH+N) in Kenya.

## Materials and methods

This qualitative study was conducted as part of the Championing Evidence Based Advocacy (CEBA) project. The project aimed to strengthen and harness the capacity of health professional associations and local champions to advocate for increased use of data in decision making, planning & resource allocation for PHC focused on RMNCAH +N in Kenya.

### Study site and participants

The study was conducted in 15 counties in Kenya: Busia, Vihiga, Migori, Kisii, Nairobi, Nakuru, West Pokot, Meru, Tharaka Nithi, Kirinyaga, Nyandarua, Mombasa, Taita Taveta, Mandera, and Wajir. County selection criteria, aimed to ensure regional representation and included counties with both high and low performance on tracer PHC/RMNCAH+N indicators during the third quarter of 2021 as captured in DHIS2 (Supplementary file 1-S1). This diverse representation enabled a nuanced understanding of the factors influencing DHIS2 data utilization across the country.

We purposely sampled and interviewed 89 key informants comprising members of the county health executive involved in health management and implementation of PHC/RMNCAH+N policies, county legislature (county assembly), representatives of health professional associations and community-level health champions. Study participants were recruited by the CEBA project county focal persons (CFPs) who were mainly paediatricians, and supported by research assistants (junior diploma-level clinician and non-clinician health workers) and community health workers.

### Data collection

All interviews were conducted by CFPs assisted by research assistants, both of whom received training in qualitative research methods, facilitated by experienced qualitative researchers at KEPRECON in March 2023. The training covered qualitative methodology, research ethics, participant recruitment, interviewing skills, reflexivity, fieldwork logistics, and transcription.

Data were collected between March, 20^th^ and May 16^th^ 2023. To prompt discussion on perceptions on DHIS2 data quality and use, participants were presented with graphs showing their county’s performance on PHC/RMNCAH+N indicators over four quarters of 2022.

They were asked to reflect on the accuracy of the trends. This was followed by a discussion on how they used DHIS2 data to support decision making and advocacy for resources to implement PHC/RMNCAH+N policies. Both CFPs and research assistants wrote reflective notes using note taking templates during and after each interview to summarise key themes and context details. Weekly debriefing sessions were held with the research project leads to provide supervision, reflect on interviewing approaches, explore emerging themes, and adjust focus for subsequent interviews. Interviews were conducted in English and Kiswahili.

Interviews were audio-recorded, transcribed, and translated verbatim into English by research assistants. 4 Quality control officers with more experience in qualitative research methods reviewed all transcripts to ensure accuracy in transcription and translation.

### Data analysis

Thematic network analysis (16) was used for data analysis, aided by Atlas.ti (version 23.2.3). Transcripts were coded inductively, based on salient themes emerging from the data and deductively, guided by the study objectives. Issues discussed by participants with respect to the study objectives within each code or set of related codes were then identified and grouped into basic themes, which were then clustered under broader organising themes. These organizing themes were subsequently synthesized into global themes, each encapsulating the main concepts conveyed within a thematic network. Several strategies were employed to enhance the credibility of the findings, including validation of the initial codebook by the technical research team (CW, DO, AA, FW) before its application to the remaining transcripts. Transcripts were independently coded by three researchers (PO, CA, MA) to enhance the reliability of the coding process. Regular debriefing sessions were held with project technical leads to discuss emerging themes, share individual viewpoints and positionalities, and mitigate potential biases in the interpretation of findings. Findings were discussed during a validation workshop that was held in October 2023 and was attended by CFPs, CHMTs from participating counties, representatives of health professional associations, champions, among other key stakeholders in health - some of whom were study participants. During this session, respective county findings were shared for sense-checking, affirmation or seeking for more relevant contextual interpretation to address any perceived researchers’ biased viewpoints.

### Ethical considerations

Ethical approval to conduct this study was obtained from AMREF Ethical and Scientific Review Committee (P1258-2022). Permission to conduct the study was also sought from the National Commission for Science, Technology, and Innovation (NACOSTI) (permit number NACOSTI P/17/1976) in Kenya. Written informed consent was sought from all participants before the interviews. Consent was reaffirmed at various points during the interviews. Several measures were taken to ensure the safety and comfort of study participants including informing participants of their right to withdraw from the study at any given point.

Participants’ confidentiality was further guaranteed through anonymisation of their identities in all research outputs.

## Results

### Characteristics of study participants

Table 1 below presents the distribution of study participants by county. To maintain anonymity and confidentiality, the names of counties have been de-identified and assigned numerical codes, and details such as participants’ age, sex, and years of experience omitted. General descriptions of participant characteristics are provided below.

**Table 1:** Distribution of key informants across counties.

| County | Participants interviewed |
| --- | --- |
| 1. | 1. Chief Health Officer-Health |
|  | 2. County Reproductive Health Coordinator |
|  | 3. Director of Curative and Preventive Services |
|  | 4. County Director of Universal Health Care |
|  | 5. Member of County Assembly |
|  | 6. Human Rights Advocate |
| 2. | 7. County Director of Health |
|  | 8. County Director of PHC Services |
|  | 9. Director of Programs |
|  | 10. Hospital Manager |
|  | 11. Head of reproductive health |
|  | 12. Member of County Assembly |
| 3. | 13. County Director of Public Health |
|  | 14. County Nutrition and Dietetics Coordinator |
|  | 15. County Reproductive Health Coordinator |
|  | 16. CEO-County Teaching and Referral Hospital |
|  | 17. NNAK representative |
|  | 18. Project Coordinator (CBO) |
|  | 19. County Assembly Health Committee Chair |
| 4. | 20. Chief Officer for Public Health |
|  | 21. County Director for Health |
|  | 22. County Nutrition Coordinator |
|  | 23. County Reproductive Health Coordinator |
|  | 24. Medical Superintendent- County Referral Hospital |
|  | 25. Programme Health Coordinator- NGO |
|  | 26. County Director for Health |
| 5. | 27. County Budget officer |
|  | 28. Member of County Assembly (party minority leader) |
|  | 29. County Reproductive Health Coordinator |
|  | 30. County Chief Officer of Health |
|  | 31. County Director of Health Services |
|  | 32. Nurse- Ripples international- NGO |
| 6. | 33. Community Based project Coordinator-NGO |
|  | 34. County Reproductive health officer |
|  | 35. Deputy Director Public health |
|  | 36. Director Health Department |
|  | 37. County Health Promotion Officer |
|  | 38. Vice chairperson health committee |
| 7. | 39. County Coordinator Reproductive health services |
|  | 40. County Director of Public Health |
|  | 41. Member of County Assembly |
| 8. | 42. County RMNCAH coordinator |
|  | 43. Chief Officer Medical Services (also acting chief officer of nutrition wellness in school feeding programme) |
|  | 44. County Child Health Coordinator |
|  | 45. County director preventive and promotive health |
|  | 46. National Nurses Association of Kenya (NNAK) representative |
|  | 47. Project Officer- Women with Disabilities group |
| 9. | 48. Chief Officer public health |
|  | 49. County Community Health Services coordinator |
|  | 50. County Nutritionist |
|  | 51. Sub County Reproductive Health Coordinator |
|  | 52. Chair health committee |
|  | 53. County Health Promotion Coordinator |
| 10. | 54. Member of County Assembly-Chair Health Committee |
|  | 55. Deputy Director of Health |
|  | 56. Member of Civil Society Organization |
|  | 57. Ministry of Health-Sub County Head |
|  | 58. Primary Health Coordinator |
|  | 59. Reproductive Health Coordinator |
| 11. | 60. Human Rights Defender |
|  | 61. MCA- Chair finance committee |
|  | 62. MCA-Chair Health Committee |
|  | 63. County Director of Health |
|  | 64. County Reproductive Health Coordinator |
|  | 65. Treasurer KCOA |
| 12. | 66. Health champion |
|  | 67. County Nursing Officer |
|  | 68. County Nutrition and Dietetics Officer |
|  | 69. Faith Based Organisation Representative |
|  | 70. MCA- Budget Committee |
|  | 71. Member of County Assembly |
| 13. | 72. MCA- Chair Budget Appropriation Committee |
|  | 73. MCA-Chair Finance Committee |
|  | 74. County Community Health Services Coordinator |
|  | 75. Representative-Health CSO coalition |
|  | 76. Deputy Director Health Administration |
|  | 77. Health Professional Association (OBS/GYN) |
| 14. | 78. County Reproductive Health Coordinator |
|  | 79. County Director of Public Health Services |
|  | 80. CSO representative - Manager Development |
|  | 81. Nurse Officer in charge- FBO |
|  | 82. MCA-Chair health committee |
|  | 83. NGO rep-Nutrition Support Officer |
| 15. | 84. CECM Health and Medical Services |
|  | 85. County Health Promotion Officer |
|  | 86. County Nutrition Coordinator |
|  | 87. County reproductive health coordinator |
|  | 88. MCA and chairperson health committee |
|  | 89. MCA and member health committee |

54 county executive members were interviewed, 22 were female and 32 were male. They comprised: Chief Health Officer-Health, Director of Curative and Preventive Services, County director preventive and promotive health, County Director of Universal Health Care, Director of Programs, hospital manager, CEO-County Teaching and Referral Hospital, Medical Superintendent-County Referral Hospital, 7 County Directors of Health Services, Deputy Director Health Administration, 2 County Health Promotion Officers, County Health Promotion Coordinators, 3 County Directors of Public Health Services, 2 Chief Officers for Public Health, Deputy Director Public health, County Budget officer, County Chief Officer of Health, County RMNCAH coordinator, Chief Officer Medical Services (acting chief officer of nutrition wellness in school feeding programme), County Child Health Coordinator, 2 County Community Health Services coordinators, Ministry of Health-Sub County Head, Primary Health Coordinator, County Director of primary health care Services, Head of reproductive health, 10 County Reproductive Health Coordinators, County Reproductive health officer, County Nursing Officer, County Executive Committee Member Health and Medical Services, 3County Nutrition and Dietetics Coordinators, County Nutrition and Dietetics Officer and County Nutritionist. Two participants had less than two years of work experience, three had between two and six years, and the remaining participants had over six years of experience. Forty-one held a university degree, five had a postgraduate qualification and eight had a post-secondary qualification that was not a university degree.

17 members of county legislatures (members of county assembly) were interviewed, 15 male and 2 female. 8 served in county health committees (6 were committee chairs and one a vice chair), 2 were chairs of the finance committee, 1 was a chair of the budget appropriation committee, 1 was a member of the budget committee. Two were female. 13 had university as their highest qualification and 4 had a post-secondary qualification that was not a university degree.

4 representatives of health professional associations, 3 were female and one male: 2 from National Nurses Association of Kenya (NNAK), one from Kenya Clinical Officers Association (KCOA) and one from the Kenya Obstetrical and Gynaecological Society OBS/GYN w14 non state actors, 8 female and 4 male, were interviewed, comprising: 3 were human rights defenders, 6 NGO representatives in charge of RMNCAH+N programmes, 2 representatives of Faith Based Organizations, 2 representatives of community-based organizations.

Even though we did not seek balanced representation of participants by sex, some patterns stood out. In one predominantly urban county (8) all key informants were female, and in another, (county 9), all but one participant was male. In one predominantly rural county (4), all key informants were male, and in another, (County 5), all but one was female. 40.4% of interviewed participants were female and 59.5% were male.

### Thematic findings

Findings are presented under two global themes (Table 2). The first global theme, use of DHIS2 data and information by CHMT members for decision making and advocacy is discussed under three organizing themes: *review of DHIS2 data*, *synthesis of DHIS2 data* and *capacity to synthesise DHIS2 data.* These themes explore socio technical influences on the use of DHIS2 generated information. Findings on the second global theme: perceptions of DHIS2 data quality are discussed under three organizing themes that explore contextual dynamics that influence perceptions of DHIS2 data quality among CHMT members, these are; (a) *Devolution* (b) *Data collection mechanisms* and (c) *Human resources to monitor indicators*.

**Table. 2.**
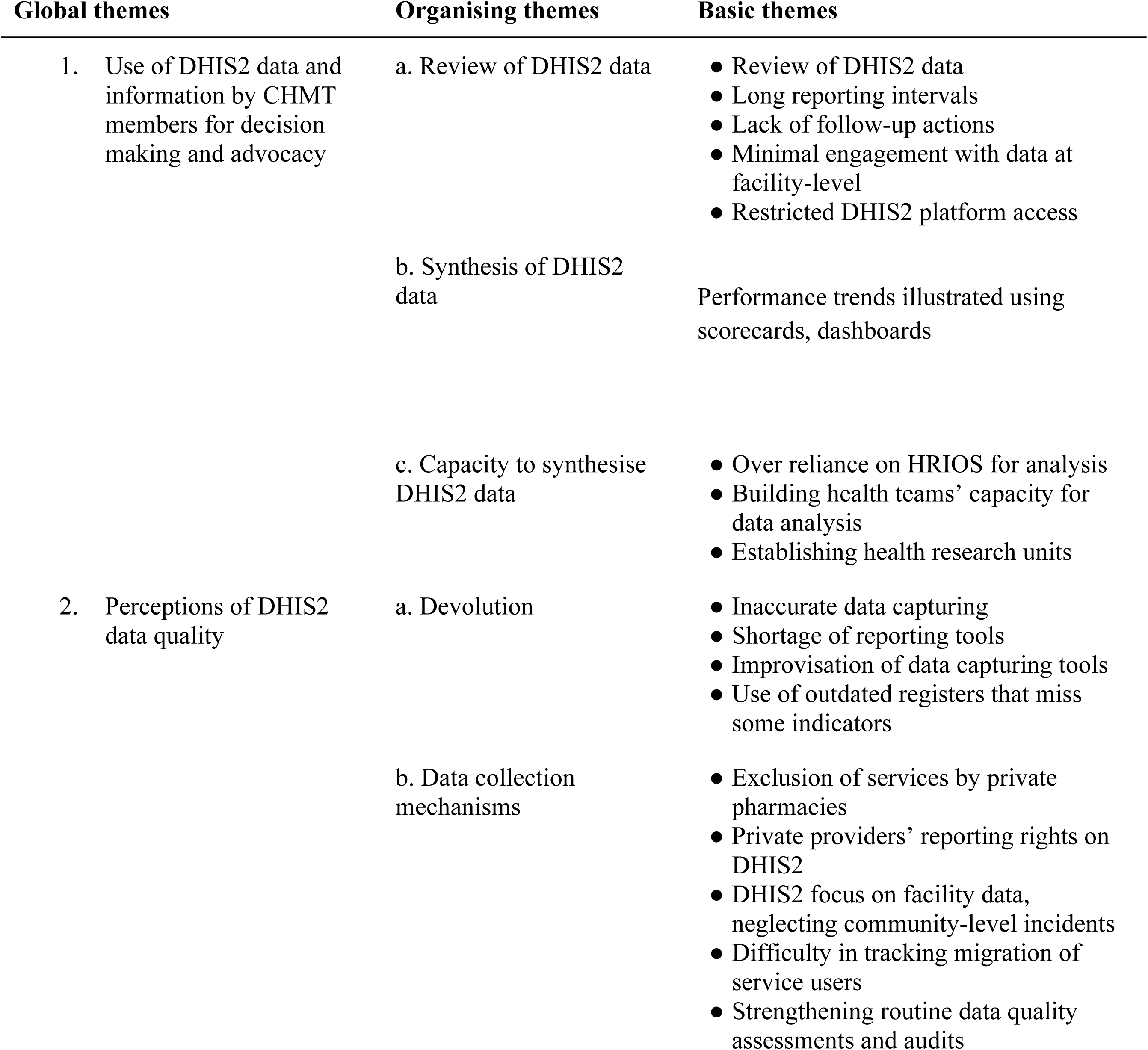
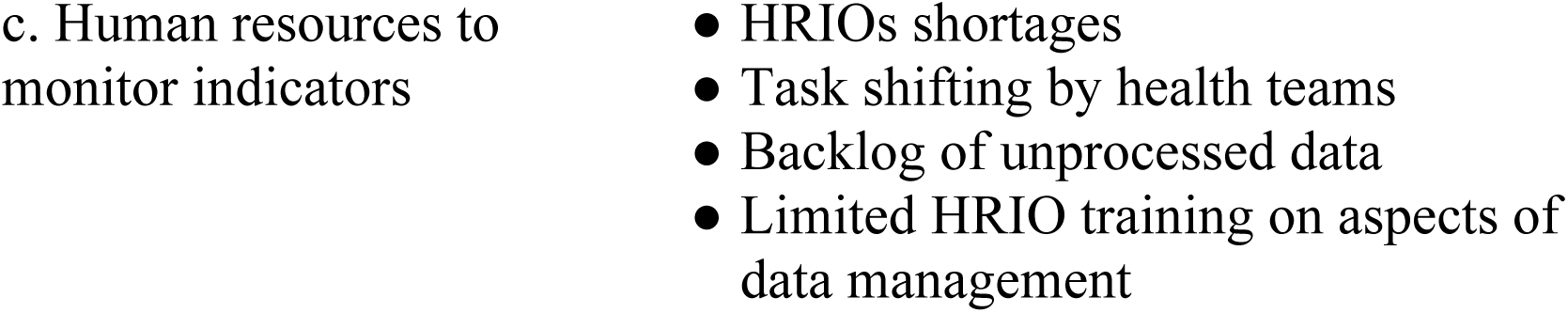
Thematic network.

### Use of DHIS2 data and information by CHMT members for decision making and advocacy

#### Review of DHIS2 data

Across all counties, CHMT members reported routinely reviewing DHIS2 generated information at the county and sub-county levels to inform decision making. They identified several gaps that undermined this practice, such as lengthy reporting intervals, lack of follow-up on actions, limited engagement with data at the facility level, and restricted access to the DHIS2 platform by stakeholders at operational levels.

CHMT members in all counties narrated that review of DHIS2 data informed resource allocation, design, and implementation of targeted interventions e.g., staff continuous medical education or community interventions to address poor indicators. In one example, elaborated in the quotation below, review of DHIS2 information revealed disparities between population and facility indicators. This suggested that residents sought healthcare services outside the county. Subsequently, the County Integrated Development Plan (CIDP) prioritised establishing a level four hospital in each sub-county.

> *So, when […] we’re in the budgeting and planning seasons, our plans are usually based on DHIS based data. And in fact, […] even the […], the current CIDP, the issue of…ah, providing level four services at the sub-county level, has actually been necessitated by this observation in our data. Where we are, […] picking that a lot of our clients are seeking services outside the county. Because our population-based data, indicates that we are doing relatively well. But when we look at service delivery data, it tells you we are not the ones who are offering those services, somebody else is. So…and we looked at the facilities, the […] sub-counties which are giving us poor indicators, and we noted there are those sub-counties that don’t have hospitals. And it has become our -- our number one priority now, to establish […] level four hospitals in each of the sub-counties. […] a serious policy, planning and budgetary […] you know decision. (Deputy Director of Health, County 10)*

CHMT members however identified the need to follow up on actions agreed upon during data review meetings in order to address poor indicators. In some counties, CHMT members also discussed the need for timely data capturing and real-time uploading (as opposed to monthly as seen in one county-13), to support timely evidence-based actions. For example, early detection of outbreaks and formulation of timely responses.

> *Health needs to get to a point that […] our health information system is real time. That we’ll be able to […] improve the responses. For instance, […], if we have an outbreak somewhere, by twenty-four hours somebody looking at the data would actually have seen the variation. But you know we up-upload at the end of the month. (Deputy Director of Health Administration County 13)*

Participants in several counties perceived limited ownership and use of DHIS2 information at the facility level. CHMT members, from Counties 2, 6, 13 and 15 noted that dialogue about indicators needs to extend beyond senior health management teams, to include facility and community-level stakeholders. The County Director of primary health care services from county 2 stated:

> *so, at the end of the month, once we’ve handed in their [facilities’] reports, I think it’s important to […] throw back the same data to the facility so that they can also see how they are doing. And so, that way, even they also consume their own data. I think there is that mechanical…uh…point of view, where you’re processing the data for […] the senior most person, but at the point where you are processing this data (background chatter) you are not also consuming the data. So, I wish we could trickle down, we– we — […], share it downwards. The information comes up, but it needs to go back down so that the facilities […] really are able to improve on the places where they’re not working well.*

An important step that was highlighted by participants to address limited engagement with data at the facility level was granting access to the DHIS2 platform to all relevant stakeholders including Health Care Providers (HCPs). For example, the sub county reproductive health coordinator from County 9 noted that currently, only the Health Records Information Officers (HRIO) had database access rights. She explained that expanding access to the DHIS2 software platform could enable HCPs to review data at their convenience, stay informed about data trends, and proactively address gaps.

> *because we don’t have rights to the DHIS, It’s only the HRIO [Health Records Information Officers] who has the rights. So, for me to analyse that data, I have to look for the HRIO. But if I had that data, and on and off I can sneak peak…they can even block it, like we cannot alter any data right on our end but it’s a read only, so that at a click of a button I can tell what is happening in the sub-county and then maybe at that point I can do something. But for me to do, like if you look at my dropout chart is not filled for this month. I have to wait for my HRIO to give me data so that I can be able to, so this one delays…*

#### Synthesis of DHIS2 data

Participants narrated that synthesis of DHIS2 data into user-friendly outputs facilitated engagement with DHIS2 data by a wide range of stakeholders and supported advocacy for increased investment in PHC/RMNCAH+N programs.

In County 7, the director of public health described that illustration of performance trends across different indicators made it easier for CHMT members to defend health budgets and secure additional resources from the County government to implement PHC/RMNCAH+N programs. She narrated:

> *You see when data is analysed like this [i.e., depicting trends as was done in documents presented to participants during the interview], it is more palatable. […] If you show anyone, they can see a trend […]. It speaks a lot too. […] I think that’s what we are trying to do now. We are doing a lot of data analytics. […] And […] we try to do that to also convince. Like for this year, when we went for budget […] defence, […] we managed to get an extra five hundred million. Hence, if you defend, you know your budget, using the data that you’ve had. […] That our indicators have really gone down, because of this and this, so you are given an additional funding.*

In counties 2 and 15, CHMT members observed that indicators, such as presenting data on teenage pregnancies by wards (the lowest electoral unit), could influence Members of County Assemblies (MCAs) to prioritise addressing poor indicators in their respective wards. In this regard, the Deputy Director Public Health from County 6 described their use of a scorecard system, with colour-coded indicators representing different performance levels for each ward.

> *we have been able to develop what we call a scorecard […] On quarterly basis, the scorecard brings out all the RMNCAH indicators -- if it is pregnancies, it is maternal, all the RMNCAH indicators. And we have been able to develop this per ward such that if you are the MCA of ward A, and we develop that scorecard, we come we invite you and members of that ward, critical stakeholders on that ward for a meeting. We might let you know your performance. And in the scorecard, we have even those […] regions which are, […] red, red, red. If your ward is red, in most of the indicators, you know you have a problem. So that has really helped us to do advocacy, and also make these critical people like the MCAs really understand that they must do something to change their red to a yellow or green.*

As is elaborated in the quotation above, scorecards served as user-friendly informative tools that enabled quick assessment of each ward’s performance by MCAs. This fostered dialogue between the county health executives and the Members of the county legislature to address poor PHC/RMNCAH+N indicators.

Participants from counties 2, 3 and 4 also recommended the implementation of colour-coded dashboards on the DHIS2 software across all health programs, for easier data visualisation of performance. They noted that dashboards, which employ colour-coding to represent performance levels on various PHC/RMNCAH+N indicators facilitate faster comprehension and enhance utilisation of data by health professionals, CHWs, health advocates, champions, and policymakers. The County Nutrition and Dietetics Coordinator from County 3 narrated:

> *I think we should come up with dashboards for specific programs in KHIS [Kenya Health Information System]. The dashboard is just a…at a glance you will be able to see what is happening. Other than figures, […]. But if there is a dashboard, you know, green means this, red means this, you know, […] yellow means this, then at a glance then you will be able to see yes, my area is not doing well, this indicator is not doing well. So, we have some areas like HIV and TB and…RH [reproductive health] having dashboards, and it is very easy to work in that area. But if we have all other programs in KHIS having their dashboard, and dashboards you know, linked properly with the data collected in KHIS, then we will be able to…to use it properly.*

The above narratives highlight that alignment between DHIS2 data outputs and end users’ preferences, capabilities and interests facilitated engagement with data by a wide range of stakeholders and supported advocacy initiatives. Notably, experiences of producing and sharing user-friendly outputs were shared in a few counties and across limited health programs, highlighting a need for replication of these practices.

#### Capacity to synthesise DHIS2 data

Despite recognising the importance of synthesising DHIS2 data for decision-making and advocacy, CHMT members in different counties expressed concern over limited existing capacity within counties to do so.

Participants from counties 6 and 10 for example suggested that enhancing the capacity of CHMT members to access, analyse, and synthesise DHIS2 data can reduce reliance on HRIOs for data analysis and interpretation.

> *First and foremost, we have very few people who know how to use DHIS. To be ascertain [precise], I think it is only HRIOs, and very few people in management, and what those in management can only do is just to look at what is entered. But we need to have several people, especially those in management who…to be taken through […] how they can analyse data, so that even if data is put into figures, how can you put them into tables and the rest and analyse and make some decision. That is one thing. Because you have only like HRIO, mostly, eh mostly the HRIO we can say are the only ones who can do several things and generate tables and graphs like these (papers shuffling) [referring to graphs presented by interviewer] and he can tell you, “This […] one you are doing badly.” Others, we can only access. (Primary Health Coordinator, County 10)*

The County Reproductive Health Coordinator from County 1 added that establishing county health research departments can further enhance the capacity of county health departments to understand dynamics that underlie poor indicators.

> *we need continuous research. In this county, we don’t have a research department […]. And that’s where we are not doing well. We are depending on external research, like the one you’re doing, but as a county we don’t have a research department. It’s [if] only County 1 can have research departments, we should be able to identify our own gaps, be it at the community level, be it at the facility level, be at the managerial level […] […] So those are the things that we want. I know that research is expensive. But that’s the […] best.*

These findings revealed a dependency on a limited pool of experts, i.e., HRIOs for DHIS2 data analysis and interpretation, highlighting the need to strengthen capabilities of other members of health teams to access, synthesise, and interpret DHIS2 data.

### Perceptions of DHIS2 data quality

Participants expressed mixed views regarding the accuracy of PHC/RMNCAH+N performance trends based on DHIS2 data that were presented to them. The majority identified data gaps that were linked to contextual factors, namely, devolution of health services, inadequacy of existing data collection mechanisms to account for patient transfers and service provision by private providers and human resource constraints.

### Devolution

CHMT members narrated that use of inappropriate reporting tools contributed to inaccurate data capturing. In 2 counties CHMT members described occasional shortages of reporting tools which led to improvisation of data capturing tools. In County 8, participants narrated that data gaps emerged due to the use of outdated registers that lacked provisions for capturing some indicators. They noted that the county had not yet developed its own data capturing registers following the devolution of health services, which had shifted this responsibility to the county level. Similarly, in County 7, participants noted that the use of outdated registers (which had since been resolved) had led to underreporting on the postnatal care indicator in previous quarters.

> *there are some things which are pulling us back […] because of the registers, you find that we have the revised registers. But […] national [government] just provided a few since health is already devolved // […] County is supposed to be now making registers for us. But it’s a challenge. […] So, you’ll find that they have gone back [to] the old registers…where we don’t have proper classification. We don’t have some of the indicators in these old registers. (County Child Health Coordinator, County 8)*

Improvisation and the use of old registers not only led to missing data on some indicators, as elaborated in the above quotation, but can also contribute to variations in monitored indicators across counties and produce incomparable data at the national level.

### Inadequate data collection mechanisms

Participants perceived gaps in DHIS2 data due to inadequacy of existing data collection mechanisms to track and patient transfers and services provided by private providers.

CHMT members in two counties indicated the actual uptake and coverage of family planning services could be higher than what was reported in the DHIS2 records, which did not account for services offered by private pharmacies, colloquially referred to as chemists.

> *if, the National [government] can work a way out whereby even the private pharmacies and chemists can be given rights to report to the national reporting platform. […] And for them to have the rights to report that facility pharmacy or whatever you’re calling it must first of all, have a KMFL (Kenya Master Health Facility) number. […] if they don’t have the KMFL number they can’t report but still there is also something else that can be done, they can be linked to a public facility. Two we tried that, and it was, and we saw that it can kind of work but sustainability was an issue. (County RMNCAH Coordinator, County 8)*

As described by the CHMT member from County 8 above, granting private providers of family planning commodities reporting rights to the DHIS2, or linking them with public health facilities to report their data is important to address underreporting of data. However, as elaborated in the above narrative, ensuring the sustainability of such strategies at the county level is challenging, indicating a need for national-level endorsement and support to implement such strategies.

In 2 other counties, participants indicated that PHC/RMNCAH+N indicator trends presented to them overstated the counties’ performance since they primarily captured health facility incidents but overlooked community incidents e.g., of malnutrition, infant and maternal mortalities, and unskilled deliveries. The County Nutrition Coordinator from County 15 indicated that health facilities in the County were underutilised.

> *The only challenge that we have with the DHIS data is the coverage. How many people come to the health facility? So, our [facility] coverage is less than 30%. So now you will realise sometimes when you have a low coverage then in the…the…. the indicators like for example how many children are malnourished will be below…way below what is actually on the ground. So, it is only the coverage.*

Participants further described migration of mothers to different locations or facilities, as seen in County 11, and their preference for accessing skilled delivery services in neighbouring counties, as seen in Counties 10 and 13. This, along with the challenge of tracing and documenting these movements, made it difficult to accurately capture data on child immunisation and skilled delivery services. The County Community Health Services Coordinator in County 13 highlighted data discrepancies that arose from capturing facility transfers by pregnant women as ‘defaulters’ in facility records, even though data from CHWs indicated no default rates.

> *what happens is that, when you go to our facilities, you will realise that we have ANC defaulters. But when you come to us in the community, we rarely have those defaulters. What happens is that a mother may decide to change a health facility.*

The challenges related to the migration of mothers between facilities/locations highlight the complexities of tracking PHC/RMNCAH+N service utilisation and the need to improve data capturing methods to account for such movements. To this end, CHMT members emphasised the need to strengthen routine data quality assessments/audits through processes such as cross-referencing data sets. They noted that these assessments should involve all stakeholders, including CHWs to interrogate and evaluate the consistency between data recorded at the community and health facilities. Participants in County 13 pointed out that inclusion of CHWs in data audits can strengthen monitoring of PHC/RMNCAH+N indicators within their communities by providing an accurate account of whether mothers have defaulted on their appointments or transferred to other facilities. In County 10, the Deputy Director of Health noted that CHWs also conduct verbal autopsies within communities which contribute to a deeper understanding of the circumstances surrounding some indicators such as child mortality.

> *we should also intensify. […] Routine Data Quality Assessments, […] moving on the ground also to compare, between what we find in registers and what we find in summary, in summary forms, and what we find in, find in, in tally sheets and stuff like that to see whether…[…] it improves on the quality of data.*

In County 9, participants highlighted an effective data capturing mechanism from the household level through CHWs to the facility level. This involved providing documentation tools to CHWs to record data at the community level. A Community Health Extension Worker (CHEW) would then compile this data into a report, which was subsequently entered into the DHIS2 system. Occasional shortages of reporting tools however led to inconsistencies between CHWs’ and facility records.

> *The tools are not adequate. […] Inadequacy of tools and therefore sometimes they [CHWs] will refer [patients] and then not capture. […] But the service was offered. […] But at the facility level we can only see an increase. […] But maybe in the community tool you may not be able to capture and relate. (County Community Health Services Coordinator, County 9)*

To mitigate the shortage of physical data capturing tools, CHMT members in Counties 5, 12, 13 and 3 recommended the adoption of electronic data capturing methods, along with the provision of necessary of necessary infrastructure, a practice that was already being implemented in County 6. The Deputy Director of Health in County 6 highlighted the successful implementation of an electronic medical records system across 12 sites. This system, which was utilised by over one thousand CHWs in the county, enabled updating of data by CHWs via mobile phones and ensured smooth integration of data collected at the community level into DHIS2.

> *We have […] I think 12 sites, where we are doing […] electrical medical records (EMR) within the county […]. We also have for our community health volunteers, we have […] slightly over…above 1000. […] currently using the digital platform, currently using phones to report on matters. Whenever they go to their houses, they report everything digitally. And this digital platform is anchoring what we call each is a national platform […] And reports are just generated now from KHIS through…it doesn’t bother them to write on paper. So, this will really help us […] We are remaining […] with other five sub counties, whose CHVs have not been digitised.*

Examples like the one above, which utilize mobile health (mHealth) technologies to support data capturing, were implemented in some sections of the county. This suggests that such interventions were potentially in their pilot phase or were donor-driven, raising questions about their sustainability.

### Human resources to monitor indicators

A recurring theme across counties was that the quality of DHIS2 data was undermined by a shortage of human resources and limited capacity among available staff to monitor indicators.

In County 10, participants noted that a shortage of HRIOs resulted in healthcare providers without adequate training in data capturing, performing multiple roles including maintaining records and report writing. In County 8, participants pointed out a substantial backlog of unprocessed data due to staff shortages. Across counties, participants thus echoed that increasing the number of available HRIOS to capture data at all levels of service delivery is crucial to enhance the quality of data uploaded into the DHIS2 system.

> *in terms of data quality, we need at least to have […] clerical officer or HI, or, or […] health information officers at most of our levels of service deliveries, […] so that we can, we can improve the quality of data that […] that […] we are uploading in our system. (Deputy Director of Health, County 10)*

Further, in County 10, participants highlighted the limited capacity of HRIOs to utilise reporting tools as a result of not receiving training on some aspects of data management. Participants in Counties 10, 13 and 15 underscored the need to strengthen the capacity of both health professionals, including HRIOs in data handling at the health facility level to improve documentation and monitoring processes.

> *There are so many reporting tools that are brought from the national or wherever, but people are not sensitised [on] what to record [on] those -- those tools. And finally, we’ll have uh…uh…data that is not the correct one, because people might be thinking what to record. So, because I think when we have uh…new tools, we need a sensitization, so that people can know exactly what am I recording, what will I be reporting. (Primary Health Coordinator, County 10)*

Overall, participants highlighted the need to strengthen stakeholders’ capabilities, through recruitment of additional HRIOs as well as the provision of regular training, including of HCPs’ on the use of data capturing tools to improve DHIS2 data accuracy.

## Discussion

We explored perceptions of DHIS2 data quality, and use of DHIS2 generated information by county-level decision makers to improve PHC/RMNCAH+N services in 15 Counties in Kenya. Findings revealed that contextual challenges, namely, counties’ differentiated access to reporting tools following the devolution of health governance, inadequacy of data collection mechanisms to trace patient transfers and account for service provision by private providers, human resource shortages and capacity constraints to capture data, contributed to DHIS2 data gaps and perceived unreliability of DHIS2 data for decision making purposes.

Regarding use of DHIS2 generated information for decision making, findings revealed that a practice of reviewing DHIS2 data - primarily at higher levels of health management, supported evidence-based decision making. Synthesis of DHIS2 data into user friendly outputs enabled a wide array of stakeholders to engage with DHIS2 data for decision making and advocacy. These practices were however undermined by limited technical capacity within counties and among health management teams to perform such synthesis and the occasional failure to implement follow up actions. Based on these findings, we identified key interventions to support use of DHIS2 data in decision-making.

The need to reform organizational culture and practices to support data ownership and use at the operational health facility level emerged as a key priority. Similar to experiences from other sub-Saharan African contexts, this study found that DHIS2 generated information was used to guide resource allocation, planning, and advocacy, (11). A significant gap that has been highlighted in this, and other studies is the limited data use at the facility level (17–19). In Kenya, previous studies have linked perceptions of limited data ownership and use at the facility level to hierarchical arrangements favouring ministry-level senior officers’ access to DHIS2 platform (19). Indeed, in the 22 health facilities in Kenya that were examined by Kihuba et al., (20) only 19% of senior managers in hospitals had access to the DHIS2 platform. This evidence underscores a need to restructure organizational culture and practices to foster value, ownership, use, and evidence-informed decisions at the health facility level. An important first step that was highlighted by participants in this study is expanding access to the DHIS2 platform by more stakeholders at the facility level.

Although we interviewed a wide range of stakeholders, only CHMT members could discuss, in depth, the use of DHIS2 evidence for decision making. County-level legislators that were interviewed had limited input about the topic, and often expressed discomfort regarding their limited understanding. Limited ability of other participant groups such as community level health champions to engage in discussions about DHIS2 data use highlights a key barrier to evidence-based decision making among key stakeholders in health. Some opportunities to facilitate engagement with DHIS2 data were identified. Narratives by CHMT members suggested that synthesis of DHIS2 data into outputs that resonated with the preferences and interests of different stakeholders supported data engagement and use across multiple users. Dashboards on the DHIS2 platform facilitated easier visualisation and comprehension of health information among CHMT members. For instance, performance trends depicted using graphs enabled CHMT to advocate for and defend health budgets. In various African countries, data charts and league tables showing districts’ indicator performance and rankings, as well as visual representation of data such as by using pie charts informed strategies for improving health services (17,18,21). These included guiding partnerships, resource allocation and advocacy efforts. In Sierra Leone, for example, stakeholder monthly review meetings to discuss charts and league tables that rank sub-districts on important indicators, awarding certificates to top performers fostered dialogue and a sense of competition, which in turn encouraged local leaders to mobilize resources and enact policies to address low uptake of services in their constituencies (21).

Despite this, our findings resonate with quantitative evidence suggesting underutilisation of DHIS2 analytical tools (17,20). CHMT members noted that dashboards existed only for some programmes, while experiences of data disaggregation by wards and the use of scorecards were only shared in a few counties. Overall, participants cited capacity limitations among CHMT members in synthesizing DHIS2 data and reliance on a limited pool of HRIOs as key barriers. Similar capacity constraints have been established in other contexts. A study that was carried out among 386 department heads from 83 health facilities in Ethiopia found that only 52% of department heads calculated achievements based on targets (22). Similar findings have been reported in Tanzania, where less than 10% of facilities in two districts conducted proper data analysis and displayed data effectively, such as plotting graphs to illustrate disease burdens (17). In Ghana, a noticeable decline in capacity from the sub-district (56.9%, 43.1%, 48.2%) to the community level (30%, 31.6% and 34.2%) was observed regarding the ability to perform tasks using the DHIS2 platform such as identifying annual targets and explaining DHIS2 data findings, plotting data, and generating monthly reports (18). Notwithstanding, a recent systematic review of interventions to address routine HIS, such as DHIS2 in LMICs, identified data analysis and display among the least addressed processes (23). Both our findings and previous literature point to a clear need for increased investment in analytical,data synthesis and visualization skills to strengthen DHIS2 data use (24). Participants in our study also noted the need to build research capacity at the county level by establishing research departments to provide research-specific technical support such as data analysis, synthesis, visualisation and interpretation to health workers.

Addressing gaps in data collection processes, focusing on strengthening coordination of public and private service providers and linkages with CHWs is key to improving DHIS2 data quality. Participants highlighted gaps in data quality relating to underreporting/overreporting of community health data. In both Malawi and Kenya, Regeru et al., (25) found that only 15% of data reported by CHWs matched consistently with reports by their supervisors. This study sheds light on some of the dynamics that underpin such discrepancies, including the migration of service users between health facilities and inadequate mechanisms to document these movements, and exclusion of community incidents e.g., of malnutrition in contexts where health facilities are underutilised. Another significant source of data gaps identified by participants in this study was the failure to integrate private service providers, such as private pharmacies, into health data reporting systems. In Kenya, private facilities account for nearly 50% of health facilities (26). Private pharmacies play a crucial role in addressing the unmet need for contraception in Kenya and other SSA contexts. Commercial drug sellers including pharmacies provide contraceptives to at least one in five young people aged 15-24 across 33 SSA countries (27,28). Our findings suggest that these contributions are inadequately captured, if any, in DHIS2 data. These gaps in DHIS2 data call for measures to establish linkages between private service providers such as pharmacies with data collection systems (29) and to strengthen data quality audits with CHWs (25).

Our findings shed light on the broader context influencing DHIS2 data quality and use. In the 15 Counties that were included in this study, participants described the impact of devolution of health governance on resources needed to support HIS. Participants described occasional shortages of reporting registers, leading to the use of outdated registers or improvisation of registers. Manya et al., (7) found that the unavailability of reporting tools across health facilities was due to the confusion over responsibilities, such as whether printing of reporting tools should be handled by the national or county governments. Participants also indicated that dependence on a limited pool of HRIOs necessitated task-shifting to other healthcare providers (HCPs) with limited training in data management and familiarity with DHIS2.

This resonates with findings by Kihuba et al. (20) who established that only 47% of recommended records officer positions were filled in the 22 facilities they examined in Kenya. Despite these findings, Kihuba et al., (20) found that HMIS departments at the facility level to be grossly underfunded. On average, they received only 3% of the total annual income from cost sharing and government grants, significantly below the policy requirement that at least 10%. While devolution of health holds promises to enhance demand for data (7), our findings highlight the need for greater investment in HIS at the County level.

### Strengths and limitations

This multicounty study shines light on the dynamics affecting DHIS2 data use at both the national and county (subnational) levels. An important contribution of this study is its highlighting both the overarching national dynamics as well as diverse practices at the county levels that shape DHIS2 data quality and use. Grounding the interpretation of findings in a sociotechnical lens offers a more critical and nuanced understanding of these dynamics. This study has several limitations. Firstly, we did not examine meeting minutes, which could have provided more insight into DHIS2 data use or non-use. Varition in participation across countries was influenced by availability and willingness of key informants to participate in the study during the data collection period. Across counties, CHMT members were more willing to participate in the study, compared to other targeted participant groups. The exclusion of these voices limits our understanding and prevents us from triangulating our findings with evidence from other stakeholders. Recruiting legislators also proved challenging, due to scheduling conflicts and concerns among legislators about the interview exposing their knowledge gaps. Even though they were assured that their responses would remain confidential, and that the purpose of the study was to gain insights rather than expose shortcomings, some declined to participate while others requested to review interview questions prior to the interview. Although 88 key informants were interviewed, legislators and health champions that were interviewed had limited input on the subject of DHIS2 data use. Findings reported thus draw mainly draw on interviews with CHMT members.

Finally, as insiders within their study contexts, CFPs were able to establish rapport with CHMT members, which facilitated the gathering of rich insights. Their insider knowledge and roles as frontline implementers of PHC/RMNCAH+N policies also enabled nuanced discussions with study participants. However, in a few cases, CFPs interviewing CHMT members (often their administrative superiors) about awareness of PHC/RMNCAH+N indicator trends and data use was perceived as interrogation or insubordination.

### Conclusions

We explored the perceptions of DHIS2 data quality and reliance for decision-making among PHC/RMNCAH+N stakeholders across 15 counties in Kenya, highlighting several contextual and structural challenges that impact data utility. Our findings indicate that while there is a strong practice of reviewing DHIS2 data at higher levels of health management, persistent data quality gaps and limitations in analytical capacity hinder its effective use, particularly at the health facility level. These challenges are compounded by differentiated access to data reporting tools post-devolution, limited integration of private service providers, and human resource constraints.

Despite these barriers, the study identified opportunities to enhance DHIS2 data use. Evidence showed that synthesizing DHIS2 data into user-friendly outputs can foster stakeholder engagement, facilitate advocacy, and support evidence-based decision-making. However, this practice remains undermined by limited technical capacity to perform data synthesis and implement follow-up actions. Addressing these limitations through investment in analytical skills, expanding data access to more stakeholders, and fostering a culture of data ownership and use at the facility level are critical to strengthening data-driven decision-making.

Targeted interventions to improve data quality are necessary, including establishing stronger coordination mechanisms between public, private, and community health actors, and enhancing data collection processes to accurately capture service delivery. Additionally, the study underscores the need for greater investment in health information systems at the county level to support routine health data collection and reporting.

Overall, this study contributes to the growing body of evidence on DHIS2 data use in sub-Saharan Africa and provides practical recommendations for improving PHC/RMNCAH+N data quality and utilisation in Kenya. Addressing the identified gaps can enable more robust use of DHIS2 data, ultimately enhancing resource allocation, planning, and service delivery for better health outcomes.

## Data Availability

The data supporting this study consists of qualitative interviews with key informants and are not publicly available due to confidentiality and ethical considerations. De-identified transcripts, summaries or excerpts may be provided upon reasonable request, subject to approval by the relevant ethics review board and in compliance with participant consent agreements.

## Acknowledgements

We express our gratitude to the Ministry of Health and participating County Governments, for granting permission to conduct this study. We also acknowledge the contributions of the research assistants who supported data collection. Finally, we sincerely appreciate the study participants for their invaluable cooperation and insights throughout the study.

PO’s contribution to this work is supported by the South African Research Chairs Initiative of the Department of Science and Technology and National Research Foundation of South Africa (Grant No. 82769).

## Authors’ contributions

FW conceptualized the study, County focal persons conducted the interviews under the coordination of PO and GO. PO undertook the analysis with support from CA and MA. PO developed the first draft of the manuscript with input from AA. All co-authors contributed to subsequent drafts and approved this version of the manuscript for publication.

## Conflict of interests, if any

The authors declare no conflict of interest.

